# Heart Rate n-Variability (HRnV) Measures for Prediction of Mortality in Sepsis Patients Presenting at the Emergency Department

**DOI:** 10.1101/2020.12.26.20248866

**Authors:** Nan Liu, Marcel Lucas Chee, Mabel Zhi Qi Foo, Jeremy Zhenwen Pong, Dagang Guo, Zhi Xiong Koh, Andrew Fu Wah Ho, Chenglin Niu, Shu-Ling Chong, Marcus Eng Hock Ong

## Abstract

**Background:** Sepsis is a potentially life threatening condition that requires prompt recognition and treatment for optimal outcomes. There is little consensus on an objective way to assess for sepsis severity and risk for mortality. In recent years, heart rate variability (HRV), a measure of the cardiac autonomic regulation derived from short electrocardiogram tracings, has been found to correlate with sepsis mortality, and its use as a prognostic variable and for risk stratification has been promising. In this paper, we present using novel heart rate n-variability (HRnV) measures for sepsis mortality risk prediction and compare against current mortality prediction scores.

**Methods:** This study was a retrospective cohort study on a convenience sample of patients presenting to the emergency department (ED) of Singapore General Hospital between September 2014 to April 2017. Patients were included in the study if they were above 21 years old, were suspected to have sepsis by their attending physician, triaged as emergency or urgent cases, and if they met two or more of the Systemic Inflammatory Response Syndrome (SIRS) criteria. Demographic and clinical variables were obtained from the electronic medical records, and HRV and novel HRnV parameters were calculated from five minute ECG tracings. Univariable analysis was conducted on variables obtained, with the primary outcome being 30-day in-hospital mortality (IHM). Variables selected through univariable analysis and stepwise selection were included in a multivariable logistic regression model, the results of which were presented using receiver operating curve (ROC) analysis.

**Results:** Of 342 patients included for final analysis, 66 (19%) met with the primary outcome. On univariable analysis, 85 out of 142 analysed HRV and HRnV parameters showed statistical difference between groups. The final multivariable logistic regression model comprised of 21 variables including four vital signs, two HRV parameters, and 15 HRnV parameters. The area under the curve (AUC) of the model was 0.86 (95% confidence interval 0.81-0.90), outperforming several established clinical scores.

**Conclusion:** The use of novel HRnV measures can provide adequate power to predictive models in the risk stratification of patients presenting to the ED with sepsis. When included in a multivariable logistic regression model, the HRnV-based model outperformed traditional risk stratification scoring systems. The HRnV measures may have potential to allow for a rapid, objective, and accurate means of patient risk stratification for sepsis severity and mortality.

## Introduction

Sepsis is a potentially life-threatening condition caused by the body’s dysregulated response to infection^1^. Every year, more than 50 million people are affected, resulting in over five million deaths worldwide^2^. Prompt recognition and treatment of sepsis has been shown to impact patient outcomes, and guidelines have been developed for its management^3^. There is, however, a need for a rapid method to grade sepsis severity and prognosticate the risk for mortality in septic patients. A quick and accurate triage tool for risk stratification of septic patients presenting at the emergency department (ED) would thus be invaluable, allowing for greater confidence to clinical decisions, and in guiding management.

Several common disease severity scoring systems that have been utilised in the ED for the prediction of sepsis mortality including the Mortality in ED Sepsis (MEDS) score^4^, quick SOFA (qSOFA)^5^, and intensive care unit (ICU)-based scores such as the Sequential Organ Failure Assessment (SOFA) score^6^, and the well-established Acute Physiology and Chronic Health Evaluation II (APACHE II) score^7^. Although these scoring systems have shown good predictive value, certain limitations have prevented their widespread adoption^8-11^. In recent years, the use of heart rate variability (HRV) measurements derived from electrocardiogram (ECG) tracings have allowed for an alternative and complementary approach for the prediction of sepsis mortality. HRV analysis measures the beat-to-beat variation between each R-R interval on an ECG tracing, and is a reflection of the autonomic regulation of the cardiovascular system^12^. Being a non-invasive tool that can be rapidly obtained even from patients who are unable to give a history, HRV has been shown to be dysregulated in sepsis^13^ and correlates well with subsequent mortality^14,15^. Indeed, scoring systems that incorporate HRV parameters among its predictors have outperformed traditional clinical indicators and established disease severity scores in predicting sepsis mortality^16-19^. The use of HRV may thus further enhance our ability to stratify for risk of sepsis mortality.

In work previously conducted by our team^20,21^, we derived additional novel heart rate n-variability (HRnV) parameters with the aim of providing enhanced prognostic information to complement traditional HRV parameters. The proposed HRnV has two measures — HR_*n*_V and HR_*n*_V_*m*_. HR_*n*_V is derived from non-overlapping R-R intervals, while HR_*n*_V_*m*_ is computed from overlapping R-R intervals. For each of the traditional HRV, HR_*n*_V, and HR_*n*_V_*m*_ measures, time domain, frequency domain, and nonlinear analysis will yield its respective set of parameters. An application of the novel HRnV variables demonstrated improved predictive ability for major adverse cardiac events among patients with chest pain presenting at the ED^21^.

In this paper, we aim to study the prognostic ability of HRnV measures alongside traditional HRV parameters in predicting the outcomes in septic patients presenting at the ED, and to compare the HRnV-based model with existing mortality prediction scores.

## Methods

### Study design and clinical setting

We conducted a retrospective cohort analysis on a convenience sample of patients presenting to Singapore General Hospital (SGH) between September 2014 to April 2017. SGH is the largest hospital in Singapore, with its ED seeing 300 to 500 patients daily. On presentation at the ED, patients are triaged according to a symptom-based Patient Acuity Category Scale (PACS). The PACS system has four levels: PACS 1 patients are critically ill, PACS 2 patients are non-ambulant but stable, PACS 3 patients are ambulant, and PACS 4 patients are non-emergency. This study was approved by the SingHealth Centralized Institutional Review Board (CIRB Ref No.: 2016/2858), with requirement for patient consent waived.

### Study population and eligibility

Patients were included in the study if they were aged 21 years and above, triaged to either PACS 1 or 2 at the ED, suspected to have sepsis as determined by their attending physician, and if they met two or more out of four Systemic Inflammatory Response Syndrome (SIRS) criteria^22,23^. The SIRS criteria (temperature >38°C or <36°C, heart rate >90 beats per minute, respiratory rate >20 breaths per minute, and total white blood cell count >12,000/mm^3^ or <4000/mm^3^) were used despite recent revisions under the Sepsis-3 consensus that recommend for sepsis screening with qSOFA score^1^. This decision was made primarily to allow for comparability with the existing literature. Additionally, subsequent validation studies have disputed the utility of qSOFA over SIRS for sepsis screening in the ED due to its poor sensitivity for septic patients^24-27^. Patients were excluded if their ECGs had non-sinus rhythm, a high noise level (>30% of the entire recording), or if they had a pacemaker or were on mechanical ventilator support.

### Data collection

Five-minute one-lead ECGs were performed on patients who met the inclusion criteria using the ZOLL X Series monitor/defibrillator (ZOLL Medical Corporation, Chelmsford, MA). Patient demographics, vital signs taken at triage, medical history, and laboratory investigations performed in the ED were retrieved from the electronic medical records. We defined the primary outcome as 30-day in-hospital mortality (IHM).

### HRnV representation and analysis

Conventional HRV analysis evaluates consecutive single RR intervals (RRIs) in ECGs. We used the two novel HRnV measures (HR_*n*_V and HR_*n*_V_*m*_) to represent beat-to-beat variations in ECG^20,21^, and have provided a visual illustration in Figure 1.

**Figure 1.**
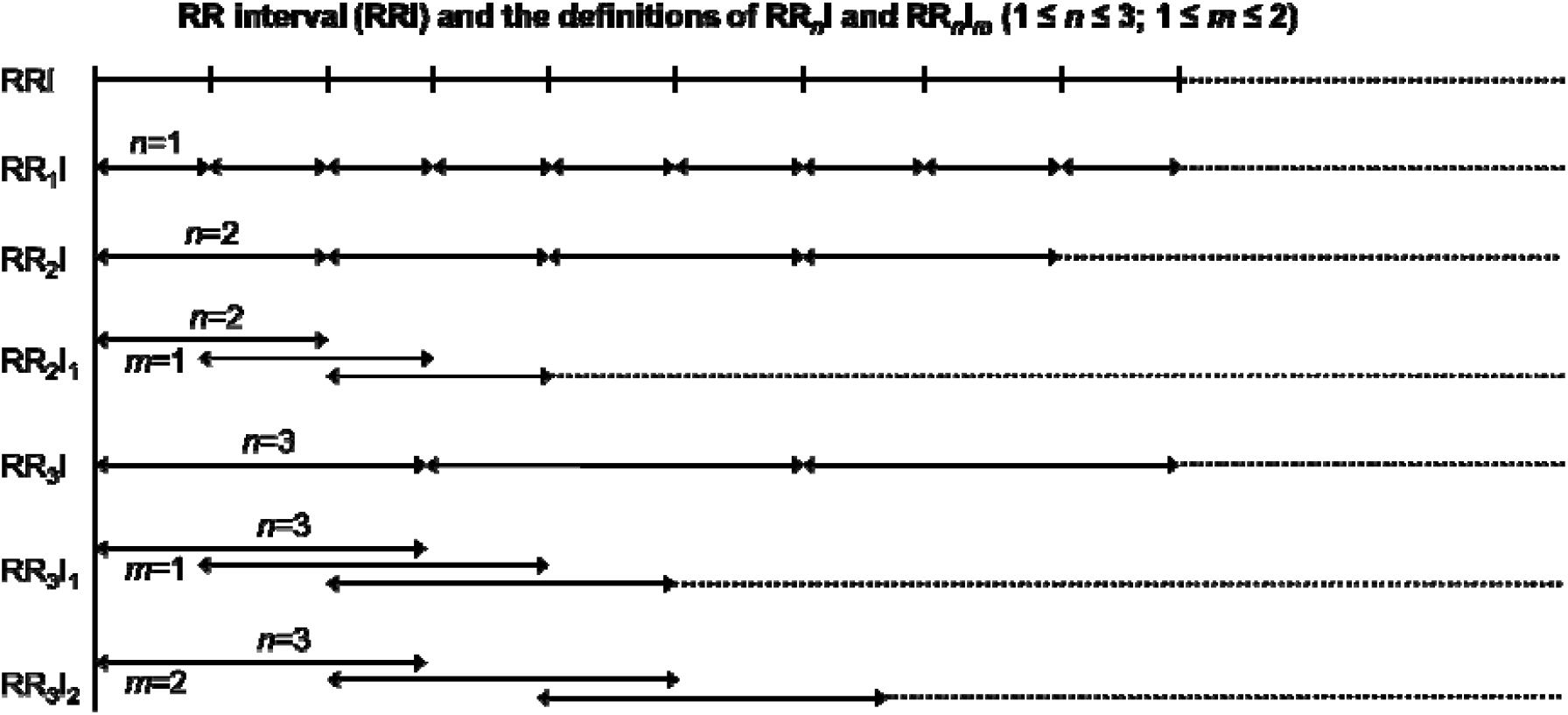
Illustration of the RR intervals (RRIs) and the definitions of RR_*n*_I and RR_*n*_I_*m*_, where 1 ≤ *n* ≤ 3, 1 ≤ *m* ≤ 2. Parameter *m* indicates the non-overlapping portion between two successive RR_*n*_I_*m*_ sequences.

To define the HR_*n*_V measure, a new type of RRI called RR_*n*_I is obtained, where *n* is an integer between 1 and *N* and *N* (number of conventional RRIs combined to form a new RR_*n*_I) is much smaller than N□ (total number of RRIs). With newly generated RR_*n*_I sequences, traditional time and frequency domains, and nonlinear analyses^28,29^ are applied to calculate HR_*n*_V parameters. In addition to conventional HRV parameters, HR_*n*_V also evaluates two newly created parameters: NN50*n* and pNN50*n*. These two parameters differ from the traditional NN50 and pNN50 parameters in that the threshold is changed from 50 ms to 50×*n* ms in describing the absolute difference between successive RR_*n*_Is.

Similarly, HR_*n*_V_*m*_ is a measure derived from RR_*n*_I_*m*_, where *m* is the number determining non-overlapping RRIs for each RR_*n*_I. When *m* = *n*, RR_*n*_I_*m*_ becomes RR_*n*_I as there are no overlapping RRIs, resulting in an upper limit of *N*-1 for *m*. Utilising all permissible combinations of *n* and *m, N*(*N*+1)/2 sets of traditional HRV, novel HR_*n*_V and HR_*n*_V_*m*_ parameters can be generated from a single RRI sequence. In our analysis, we set the upper limit of *N* as 3, due to the relatively short duration of collected ECG samples. As a result, one set of HRV parameters, two sets of HR_*n*_V (HR_2_V and HR_3_V) parameters, and three sets of HR_*n*_V_*m*_ (HR_2_V_1_, HR_3_V_1_, and HR_3_V_2_) parameters can be calculated. The HRnV-Calc software suite (https://github.com/nliulab/HRnV) was used for calculating the HRV and HRnV parameters, in which the functions from PhysioNet Cardiovascular Signal Toolbox^30^ were performed for ECG signal processing.

### Statistical analysis

Categorical variables were compared between patients who did and did not meet the primary outcome (30-day IHM) using χ^2^ test or Fisher’s exact test where appropriate. Continuous variables were checked for normality with the Kolmogorov-Smirnov test. Subsequently, normally distributed variables were presented as mean and standard deviation (SD) and were compared with independent two-tailed *t* test between groups, while non-normally distributed variables were presented as median and interquartile range (IQR; 25^th^ to 75^th^ percentiles) and compared using the Mann-Whitney U test.

Univariable regression analysis was conducted on traditional HRV parameters, novel HRnV parameters and demographic and clinical variables. Each variable was evaluated as an individual predictor of the primary outcome (30-day IHM) using binary logistic regression with odds ratio (OR), 95% confidence interval (CI) and p-value reported. For multivariable regression analysis, we adjusted for age, temperature, systolic blood pressure, heart rate, and Glasgow Coma Scale (GCS) as these variables were either shown to be significant predictors of sepsis mortality in previous literature^15,31-33^, or are included in well-established sepsis scoring systems such as the National Early Warning Score (NEWS)^34^, Modified Early Warning Score (MEWS)^35^, MEDS, qSOFA, or APACHE II. HRV and HRnV parameters were included in the multivariable analysis if they achieved p<0.2 in the univariable analysis. Included variables were then checked for collinearity using Pearson’s R correlation. For each collinear pair, the variable with the higher p-value on univariate analysis was eliminated until no collinear pairs remained. The remaining variables were then fed into a backward stepwise multivariable logistic regression model which used p<0.1 as an endpoint. We took statistical significance at p-value<0.05. A receiver operating characteristic (ROC) curve was plotted to assess the predictive ability of the multivariable regression model and compared against other established disease scoring systems on their area under the curve (AUC).

Missing data were addressed by median imputation, in consideration of the low proportion of missing data (<0.3%) for each variable, the nature of variables, and recommendations for missing data in clinical trials^36^. There were three missing observations for which the median value was imputed; one patient had unknown medical history of cancer, and another patient was missing both initial and worst qSOFA scores.

All statistical analyses were carried out using Python version 3.8.0 (Python Software Foundation, Delaware, USA) using the SciPy library (version 1.3.1). Regression models were built using the StatsModels library (version 0.10.2) and scikit-learn library (version 0.22).

## Results

### Patient recruitment

Figure 2 presents the patient recruitment flowchart. Of the 659 patients that were initially recruited, 190 patients did not meet the SIRS criteria, and 127 patients had inapplicable ECG readings. A total of 342 patients were included for analysis and classified depending on whether they met the primary outcome of 30-day IHM (n=66, 19%), or did not meet the primary outcome (n=276, 81%).

**Figure 2.**
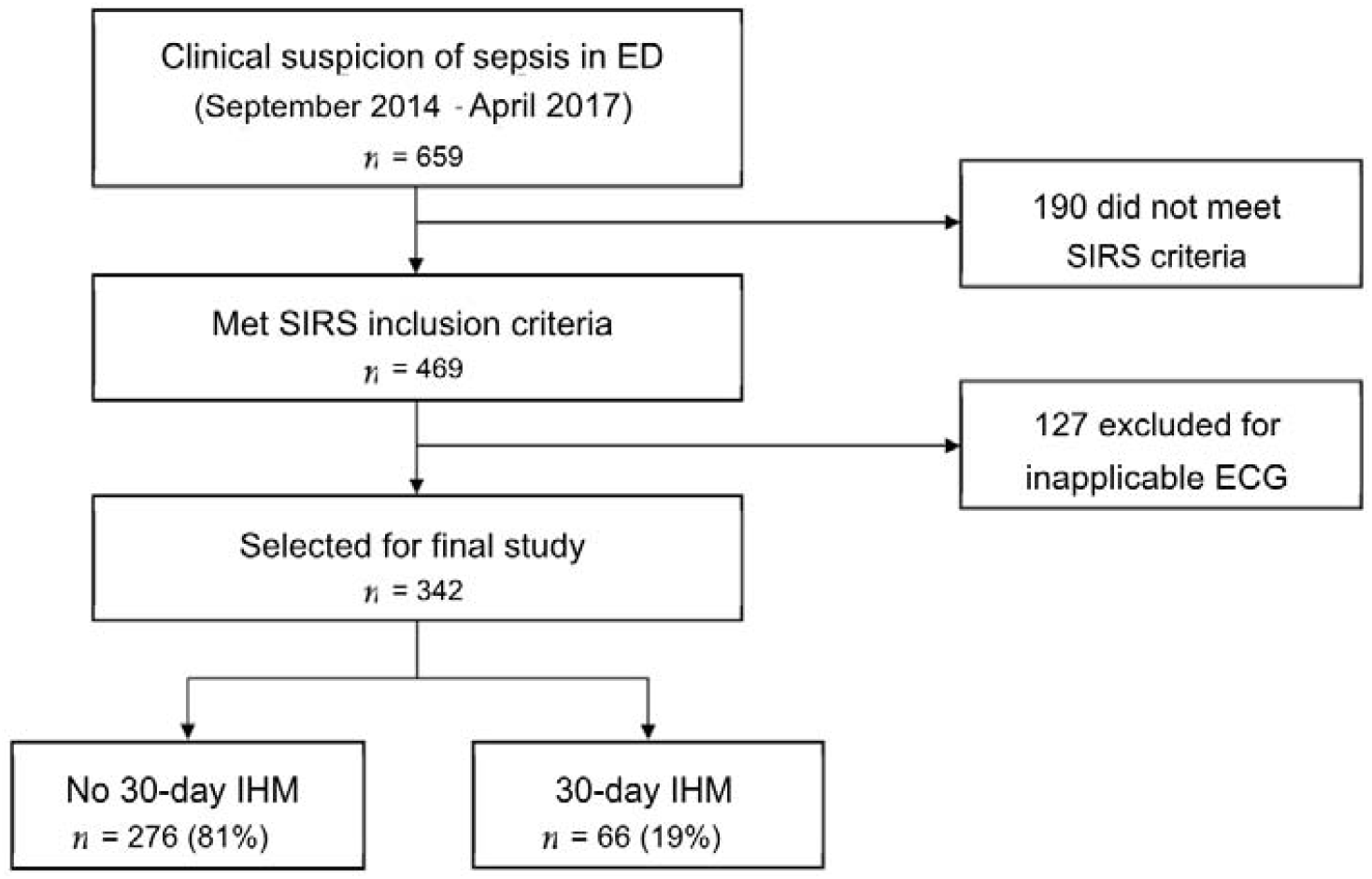
Patient recruitment flowchart. ECG: Electrocardiogram; ED: emergency department; IHM: in-hospital mortality; SIRS: systemic inflammatory response syndrome.

### Baseline characteristics and clinical parameters

Table 1 illustrates baseline characteristics and clinical parameters of patients who met and did not meet with 30-day IHM. Patients who met with 30-day IHM were older and presented with higher respiratory rates but lower temperatures, systolic blood pressures (SBP) and Glagow Coma Scale (GCS) scores, when compared to patients who did not meet with 30-day IHM. The worst recorded values of respiratory rate, GCS, and SBP during each patient’s ED stay were also significantly more abnormal in patients that met with 30-day IHM. Difference in disposition from the ED was significant, with a larger proportion of patients who eventually met with 30-day IHM requiring admission to the ICU as compared to patients who did not meet with 30-day IHM (16.7% vs 4.3%, p=0.001). Additionally, a larger proportion of patients who met with 30-day IHM had a respiratory source of infection (45.5% vs 27.2%, p=0.006) while a smaller proportion had a source of infection originating from the urinary tract (7.6% vs 25.7%, p=0.003) when compared to patients who did not meet with 30-day IHM. No significant differences were detected in gender, PACS status, ethnicity, or medical history between both groups.

**Table 1.** Baseline characteristics and clinical parameters.

| Variable | No 30-day IHM<br>(n=276) | 30-day IHM<br>(n=66) | P-value |
| --- | --- | --- | --- |
| Age, mean (SD) | 65.8 (16.1) | 73.2 (14.8) | 0.001* |
| Male gender, n (%) | 144.0 (52.2) | 30.0 (45.5) | 0.399 |
| Triaged to high acuity (PACS1), n (%) | 254.0 (92.0) | 64.0 (97.0) | 0.19 |
| SIRS criteria met, n (%) | 250.0 (90.6) | 58.0 (87.9) | 0.667 |
| **Race, n (%)** |  |  | 0.946 |
| Chinese | 203.0 (73.6) | 49.0 (74.2) | 0.967 |
| Malay | 40.0 (14.5) | 8.0 (12.1) | 0.763 |
| Indian | 21.0 (7.6) | 6.0 (9.1) | 0.883 |
| Other | 12.0 (4.3) | 3.0 (4.5) | 1 |
| **Disposition from ED, n (%)** |  |  | 0.001* |
| Intensive care unit | 12.0 (4.3) | 11.0 (16.7) | 0.001* |
| high-dependency unit | 28.0 (10.1) | 3.0 (4.5) | 0.231 |
| General ward | 236.0 (85.5) | 52.0 (78.8) | 0.247 |
| **Medical history, n (%)** |  |  |  |
| Ischemic heart disease | 73.0 (26.4) | 21.0 (31.8) | 0.469 |
| Diabetes | 111.0 (40.2) | 24.0 (36.4) | 0.663 |
| Hypertension | 156.0 (56.5) | 35.0 (53.0) | 0.708 |
| Cancer | 79.0 (28.6) | 23.0 (34.8) | 0.399 |
| Serious infection | 117.0 (42.4) | 28.0 (42.4) | 0.894 |
| **Source of infection, n (%)** |  |  |  |
| Respiratory | 75.0 (27.2) | 30.0 (45.5) | 0.006* |
| Urinary tract | 71.0 (25.7) | 5.0 (7.6) | 0.003* |
| Gastrointestinal | 18.0 (6.5) | 4.0 (6.1) | 1 |
| Musculoskeletal | 11.0 (4.0) | 3.0 (4.5) | 0.738 |
| Hepatobiliary | 20.0 (7.2) | 0.0 (0.0) | 0.018* |
| Peritoneum | 3.0 (1.1) | 2.0 (3.0) | 0.248 |
| Skin | 3.0 (1.1) | 0.0 (0.0) | 1 |
| Line | 7.0 (2.5) | 0.0 (0.0) | 0.354 |
| Cardiac | 7.0 (2.5) | 2.0 (3.0) | 0.686 |
| Central nervous system | 1.0 (0.4) | 0.0 (0.0) | 1 |
| Unknown | 23.0 (8.3) | 12.0 (18.2) | 0.032* |
| No infection | 37.0 (13.4) | 8.0 (12.1) | 0.94 |
| **Vital sign predictors, mean (SD) or median (IQR)** |  |  |  |
| Heart rate, bpm | 114.2 (23.3) | 112.7 (26.0) | 0.652 |
| White blood cell count | 14.0 (7.7) | 13.0 (9.6) | 0.36 |
| Diastolic BP, mmHg | 63.0 (19.5) | 59.7 (17.4) | 0.213 |
| Temperature, °C | 38.1 (37.1-38.8) | 37.2 (36.3-38.0) | <0.001* |
| Respiratory rate, bpm | 19.0 (18.0-21.0) | 22.0 (19.0-25.0) | <0.001* |
| Respiratory rate (worst), bpm | 21.0 (19.8-24.0) | 26.0 (22.0-30.0) | <0.001* |
| Systolic BP, mmHg | 109.0 (86.0-139.0) | 101.0 (78.0-118.5) | 0.012* |
| Systolic BP (worst), mmHg | 90.0 (77.0-109.2) | 78.0 (63.2-94.8) | <0.001* |
| GCS (3-15) | 13.4 (3.0) | 11.7 (4.1) | 0.001* |
| Worst GCS (3-15) | 13.4 (3.1) | 11.7 (4.1) | 0.002* |

\*p<0.05
IHM: in-hospital mortality; SD: standard deviation; IQR: interquartile range; BP: blood pressure; GCS: Glasgow Coma Scale; ED: emergency department

### HRV and HRnV parameter description and univariable analysis

Table 2 presents the descriptive analysis of HRV and HRnV parameters. In this study, *N* was set as 3 and HR_2_V, HR_2_V_1_, HR_3_V, HR_3_V_1_ and HR_3_V_2_ parameters were calculated. Among time domain parameters such as mean NN and SDNN, HR_*n*_V and HR_*n*_V_*m*_ values are generally directly proportional to *n* and increase when *n* increases. HR_2_V SampEn and HR_3_V SampEn were considerably larger compared to SampEn parameters of HRV, HR_2_V_1_, HR_3_V_1_, and HR_3_V_2_. This was due to insufficient data points since our ECG recordings were only five minutes long. HR_2_V_1_, HR_3_V_1_ and HR_3_V_2_ did not encounter this limitation as more data points were available from calculation using overlapping RR_*n*_I_*m*_ sequences^21^.

**Table 2.**
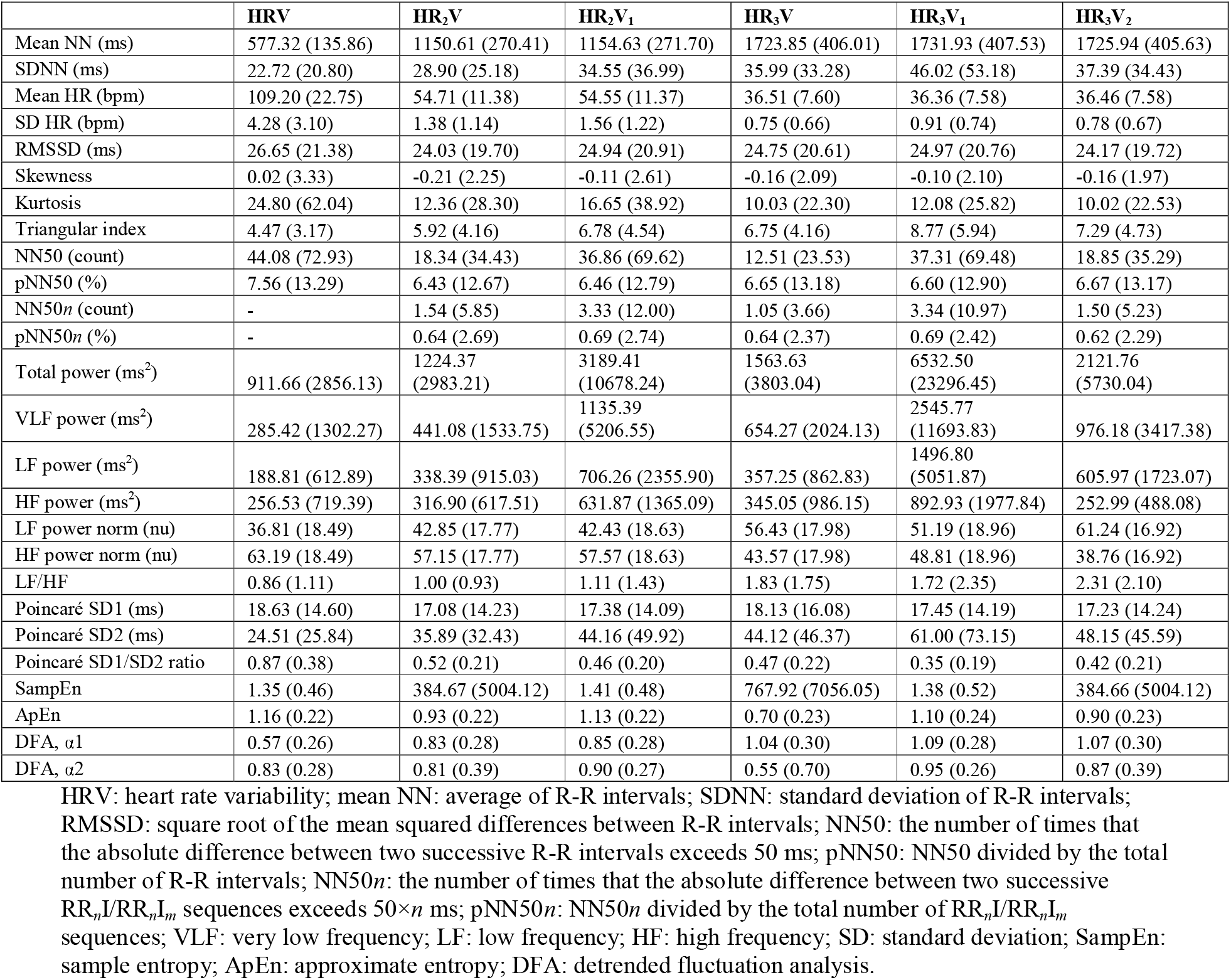
Descriptive analyses of heart rate variability (HRV) and heart rate n-variability (HRnV) parameters

Table 3 shows the results of univariable analysis of HRV and HRnV parameters. Of 142 HRV and HRnV parameters, 85 were significantly different between the two outcome groups. Specifically, 14 HRV, 14 HR_2_V, 16 HR_2_V_1_, 11 HR_3_V, 16 HR_3_V_1_, and 14 HR_3_V_2_ parameters were statistically significant. In at least four out of six HRnV measures, RMSSD, kurtosis, NN50, pNN50, NN50*n*, pNN50*n*, HF power, HF power norm, Poincare SD1, and Poincare SD1/SD2 were significantly higher, while LF power norm and DFA α2 were significantly lower in patients who met the primary outcome compared to those who did not. Additionally, VLF power and DFA α1 were not significant in HRV analysis but were statistically significant in several HRnV measures.

**Table 3.** Univariable analysis of HRV and HRnV parameters

|  | HRV |  | HR<sub>2</sub>V |  | HR<sub>3</sub>V |  |
| --- | --- | --- | --- | --- | --- | --- |
|  | OR (95% CI) | p | OR (95% CI) | P | OR (95% CI) | p |
| Mean NN | 1.000 (0.998 to 1.002) | 0.903 | 1.000 (0.999 to 1.001) | 0.964 | 1.000 (0.999 to 1.001) | 0.978 |
| SDNN | 1.021 (1.008 to 1.035) | 0.002* | 1.011 (1.002 to 1.020) | 0.021* | 1.007 (1.000 to 1.014) | 0.065 |
| RMSSD | 1.021 (1.009 to 1.032) | <0.001* | 1.020 (1.008 to 1.032) | 0.001* | 1.019 (1.007 to 1.030) | 0.002* |
| Skewness | 1.019 (0.943 to 1.102) | 0.632 | 0.982 (0.873 to 1.105) | 0.762 | 1.046 (0.916 to 1.193) | 0.508 |
| Kurtosis | 1.003 (0.999 to 1.007) | 0.12 | 1.007 (0.999 to 1.015) | 0.089 | 1.010 (0.999 to 1.020) | 0.063 |
| Triangular index | 1.044 (0.967 to 1.128) | 0.271 | 1.029 (0.970 to 1.091) | 0.342 | 1.014 (0.952 to 1.079) | 0.668 |
| NN50 | 1.004 (1.001 to 1.007) | 0.016* | 1.007 (1.001 to 1.014) | 0.034* | 1.010 (1.000 to 1.020) | 0.048* |
| pNN50 | 1.024 (1.006 to 1.042) | 0.008* | 1.023 (1.004 to 1.041) | 0.016* | 1.019 (1.002 to 1.037) | 0.032* |
| NN50n | - | - | 1.055 (1.013 to 1.099) | 0.011* | 1.087 (1.023 to 1.155) | 0.007* |
| pNN50n | - | - | 1.122 (1.025 to 1.228) | 0.013* | 1.136 (1.034 to 1.248) | 0.008* |
| Total power | 1.000 (1.000 to 1.000) | 0.007* | 1.000 (1.000 to 1.000) | 0.015* | 1.000 (1.000 to 1.000) | 0.056 |
| VLF power | 1.000 (1.000 to 1.001) | 0.079 | 1.000 (1.000 to 1.000) | 0.058 | 1.000 (1.000 to 1.000) | 0.039* |
| LF power | 1.001 (1.000 to 1.001) | 0.004* | 1.000 (1.000 to 1.001) | 0.021* | 1.000 (1.000 to 1.001) | 0.018* |
| HF power | 1.001 (1.000 to 1.001) | 0.004* | 1.001 (1.000 to 1.001) | 0.001* | 1.000 (1.000 to 1.000) | 0.268 |
| LF power norm | 0.978 (0.962 to 0.995) | 0.009* | 0.987 (0.972 to 1.002) | 0.095 | 0.986 (0.972 to 1.001) | 0.059 |
| HF power norm | 1.022 (1.005 to 1.039) | 0.009* | 1.013 (0.998 to 1.029) | 0.095 | 1.014 (0.999 to 1.029) | 0.059 |
| LF/HF | 0.682 (0.457 to 1.017) | 0.061 | 0.856 (0.618 to 1.187) | 0.352 | 0.960 (0.810 to 1.138) | 0.637 |
| Poincaré SD1 | 1.027 (1.010 to 1.044) | 0.001* | 1.028 (1.011 to 1.045) | 0.001* | 1.021 (1.007 to 1.037) | 0.004* |
| Poincaré SD2 | 1.013 (1.002 to 1.024) | 0.018* | 1.007 (1.000 to 1.014) | 0.066 | 1.003 (0.998 to 1.008) | 0.235 |
| Poincaré SD1/SD2 | 2.346 (1.160 to 4.745) | 0.018* | 8.160 (2.164 to 30.773) | 0.002* | 8.285 (2.541 to 27.017) | <0.001* |
| SampEn | 0.965 (0.534 to 1.741) | 0.905 | 1.000 (1.000 to 1.000) | 0.31 | 1.000 (1.000 to 1.000) | 0.15 |
| ApEn | 0.269 (0.083 to 0.879) | 0.03* | 0.261 (0.079 to 0.862) | 0.028* | 0.729 (0.232 to 2.288) | 0.588 |
| DFA, $\alpha_1$ | 0.704 (0.243 to 2.038) | 0.518 | 0.342 (0.123 to 0.955) | 0.041* | 0.213 (0.077 to 0.590) | 0.003* |
| DFA, $\alpha_2$ | 0.137 (0.047 to 0.400) | <0.001* | 0.352 (0.177 to 0.699) | 0.003* | 0.631 (0.433 to 0.918) | 0.016* |
|  | HR<sub>2</sub>V<sub>1</sub> |  | HR<sub>3</sub>V<sub>1</sub> |  | HR<sub>3</sub>V<sub>2</sub> |  |
|  | OR (95% CI) | p | OR (95% CI) | p | OR (95% CI) | p |
| Mean NN | 1.000 (0.999 to 1.001) | 0.905 | 1.000 (0.999 to 1.001) | 0.906 | 1.000 (0.999 to 1.001) | 0.965 |
| SDNN | 1.010 (1.003 to 1.018) | 0.008* | 1.006 (1.001 to 1.012) | 0.018* | 1.007 (1.000 to 1.013) | 0.053 |
| RMSSD | 1.022 (1.011 to 1.034) | <0.001* | 1.021 (1.010 to 1.033) | <0.001* | 1.019 (1.007 to 1.032) | 0.002* |
| Skewness | 0.934 (0.842 to 1.036) | 0.198 | 0.909 (0.799 to 1.032) | 0.141 | 0.954 (0.836 to 1.089) | 0.485 |
| Kurtosis | 1.007 (1.001 to 1.013) | 0.022* | 1.010 (1.001 to 1.018) | 0.03* | 1.009 (0.999 to 1.019) | 0.092 |
| Triangular index | 1.027 (0.972 to 1.085) | 0.34 | 1.012 (0.969 to 1.057) | 0.586 | 1.009 (0.954 to 1.066) | 0.763 |
| NN50 | 1.004 (1.001 to 1.007) | 0.022* | 1.004 (1.000 to 1.007) | 0.034* | 1.007 (1.000 to 1.014) | 0.043* |
| pNN50 | 1.024 (1.006 to 1.042) | 0.009* | 1.022 (1.004 to 1.040) | 0.017* | 1.020 (1.002 to 1.038) | 0.027* |
| NN50n | 1.026 (1.006 to 1.045) | 0.009* | 1.028 (1.007 to 1.049) | 0.007* | 1.064 (1.020 to 1.110) | 0.004* |
| pNN50n | 1.120 (1.028 to 1.220) | 0.01* | 1.138 (1.036 to 1.249) | 0.007* | 1.151 (1.044 to 1.270) | 0.005* |
| Total power | 1.000 (1.000 to 1.000) | 0.015* | 1.000 (1.000 to 1.000) | 0.022* | 1.000 (1.000 to 1.000) | 0.031* |
| VLF power | 1.000 (1.000 to 1.000) | 0.08 | 1.000 (1.000 to 1.000) | 0.08 | 1.000 (1.000 to 1.000) | 0.059 |
| LF power | 1.000 (1.000 to 1.000) | 0.005* | 1.000 (1.000 to 1.000) | 0.005* | 1.000 (1.000 to 1.000) | 0.026* |
| HF power | 1.000 (1.000 to 1.000) | 0.002* | 1.000 (1.000 to 1.000) | 0.001* | 1.001 (1.000 to 1.001) | 0.001* |
| LF power norm | 0.980 (0.965 to 0.996) | 0.013* | 0.979 (0.965 to 0.994) | 0.006* | 0.979 (0.964 to 0.995) | 0.009* |
| HF power norm | 1.020 (1.004 to 1.037) | 0.013* | 1.021 (1.006 to 1.037) | 0.006* | 1.021 (1.005 to 1.038) | 0.009* |
| LF/HF | 0.769 (0.570 to 1.037) | 0.085 | 0.883 (0.744 to 1.047) | 0.153 | 0.884 (0.752 to 1.040) | 0.137 |
| Poincaré SD1 | 1.029 (1.012 to 1.047) | 0.001* | 1.028 (1.011 to 1.045) | 0.001* | 1.027 (1.010 to 1.044) | 0.002* |
| Poincaré SD2 | 1.006 (1.000 to 1.011) | 0.034* | 1.004 (1.000 to 1.007) | 0.048* | 1.004 (0.999 to 1.009) | 0.129 |
| Poincaré SD1/SD2 | 7.893 (2.085 to 29.879) | 0.002* | 7.405 (1.867 to 29.365) | 0.004* | 7.610 (2.222 to 26.064) | 0.001* |
| SampEn | 0.830 (0.471 to 1.461) | 0.518 | 0.809 (0.480 to 1.364) | 0.427 | 1.000 (1.000 to 1.000) | 0.31 |
| ApEn | 0.382 (0.122 to 1.192) | 0.097 | 0.728 (0.244 to 2.170) | 0.569 | 0.544 (0.175 to 1.693) | 0.293 |
| DFA, $\alpha_1$ | 0.480 (0.171 to 1.350) | 0.164 | 0.471 (0.173 to 1.284) | 0.141 | 0.380 (0.146 to 0.990) | 0.048* |
| DFA, $\alpha_2$ | 0.141 (0.047 to 0.423) | <0.001* | 0.146 (0.048 to 0.448) | 0.001* | 0.384 (0.194 to 0.760) | 0.006* |

\* $p < 0.05$ . HRV: heart rate variability; OR: odds ratio; CI: confidence interval; mean NN: average of R-R intervals; SDNN: standard deviation of R-R intervals; RMSSD: square root of the mean squared differences between R-R intervals; NN50: the number of times that the absolute difference between two successive R-R intervals exceeds 50 ms; pNN50: NN50 divided by the total number of R-R intervals; NN50n: the number of times that the absolute difference between 2 successive $RR_n/RR_nI_m$ sequences exceeds $50 \times n$ ms; pNN50n: NN50n divided by the total number of $RR_n/RR_nI_m$ sequences; VLF: very low frequency; LF: low frequency; HF: high frequency; SD: standard deviation; SampEn: sample entropy; ApEn: approximate entropy; DFA: detrended fluctuation analysis.

Overall, six baseline characteristics (age and vital signs at triage including temperature, respiratory rate, SpO_2_, SBP and GCS), 17 HRV parameters, and 96 HRnV parameters had p<0.2 on univariable analysis. After collinearity assessment, the remaining 87 variables were entered into a stepwise-selection regression model.

### Multivariable analysis and ROC analysis

Table 4 presents the multivariable analysis of variables found to be significantly different on univariable analysis. A total of 21 out of 87 variables were selected through stepwise selection. Of the 21 variables, 16 showed p<0.05. These include vital signs such as respiratory rate (OR=1.168; 95% CI 1.085-1.257; p<0.001), SBP (OR=0.978; 95% CI 0.966-0.990; p=0.001), SpO_2_ (OR=0.892; 95% CI 0.838-0.950; p=<0.001), and GCS (OR=0.845; 95% CI 0.769-0.929; p=0.001), and HRnV measures such as HR_2_V_1_ NN50 (OR=0.808; 95% CI 0.682-0.958; p=0.014), HR_2_V pNN50 (OR=0.290; 95% CI 0.115-0.732; p=0.009), HR_2_V_1_ pNN50 (OR=5.700; 95% CI 1.784-18.213; p=0.003), HR_2_V ApEn (OR=0.106; 95% CI 0.013-0.877; p=0.037) and several HR_3_V_1_ and HR_3_V_2_ parameters which demonstrated strong predictive power in assessing the risk for 30-day IHM. The final multivariable predictive model consisted of four vital signs, two traditional HRV parameters, and 15 novel HRnV parameters. Hereafter, we refer to this model as the HRnV model.

**Table 4:** Multivariable analysis

| Variables | Adjusted Odds Ratio (95% CI) | p-value |
| --- | --- | --- |
| Vital signs |  |  |
| Respiratory rate | 1.168 (1.085 to 1.257) | <0.001* |
| Systolic blood pressure | 0.978 (0.966 to 0.990) | 0.001* |
| Glasgow coma scale | 0.845 (0.769 to 0.929) | 0.001* |
| SpO<sub>2</sub> | 0.892 (0.838 to 0.950) | <0.001* |
| HRV parameters |  |  |
| HRV total power | 1.000 (1.000 to 1.001) | 0.019* |
| HRV Poincare SD1 | 0.948 (0.893 to 1.007) | 0.081 |
| HRnV parameters |  |  |
| HR<sub>2</sub>V NN50 | 1.270 (0.967 to 1.667) | 0.085 |
| HR<sub>2</sub>V HF power | 1.002 (1.000 to 1.003) | 0.076 |
| HR<sub>2</sub>V pNN50 | 0.290 (0.115 to 0.732) | 0.009* |
| HR<sub>2</sub>V ApEn | 0.106 (0.013 to 0.877) | 0.037* |
| HR<sub>2</sub>V<sub>1</sub> NN50 | 0.808 (0.682 to 0.958) | 0.014* |
| HR<sub>2</sub>V<sub>1</sub> pNN50 | 5.700 (1.784 to 18.213) | 0.003* |
| HR<sub>2</sub>V<sub>1</sub> LF/HF | 0.251 (0.071 to 0.880) | 0.031* |
| HR<sub>3</sub>V pNN50 | 1.229 (0.988 to 1.528) | 0.064 |
| HR<sub>3</sub>V<sub>1</sub> NN50 | 1.098 (0.999 to 1.206) | 0.053 |
| HR<sub>3</sub>V<sub>1</sub> LF/HF | 2.241 (1.252 to 4.013) | 0.007* |
| HR<sub>3</sub>V<sub>1</sub> pNN50 | 0.473 (0.247 to 0.906) | 0.024* |
| HR<sub>3</sub>V<sub>1</sub> NN50<sub>n</sub> | 0.794 (0.664 to 0.950) | 0.012* |
| HR<sub>3</sub>V<sub>1</sub> HF power norm | 1.093 (1.044 to 1.144) | <0.001* |
| HR<sub>3</sub>V<sub>1</sub> DFA α1 | 14.189 (1.009 to 199.510) | 0.049* |
| HR<sub>3</sub>V<sub>2</sub> NN50<sub>n</sub> | 1.713 (1.202 to 2.443) | 0.003* |

\*p<0.05
HRV: heart rate variability; CI: confidence interval; NN50: the number of times that the absolute difference between two successive R-R intervals exceeds 50 ms; pNN50: NN50 divided by the total number of R-R intervals; NN50<sub>n</sub>: the number of times that the absolute difference between 2 successive RR<sub>n</sub>/RR<sub>n</sub>I<sub>m</sub> sequences exceeds 50×n ms; LF: low frequency; HF: high frequency; SD: standard deviation; ApEn: approximate entropy; DFA: detrended fluctuation analysis.

A ROC curve was plotted for assessment of the HRnV model and compared against established disease severity scoring systems for the prediction of 30-day IHM in patients presenting to the ED with sepsis (Figure 3). The AUC of the HRnV model was 0.86 (95% CI: 0.81-0.90), outperforming the AUC of NEWS 0.71 (95% CI: 0.64-0.78), MEWS 0.60 (95% CI: 0.53-0.67), MEDS 0.85 (95% CI: 0.81-0.90), SOFA 0.71 (95% CI: 0.64-0.78), APACHE II 0.74 (95% CI: 0.68-0.80), and the patient’s worst qSOFA value 0.72 (95% CI: 0.65-0.79).

**Figure 3.**
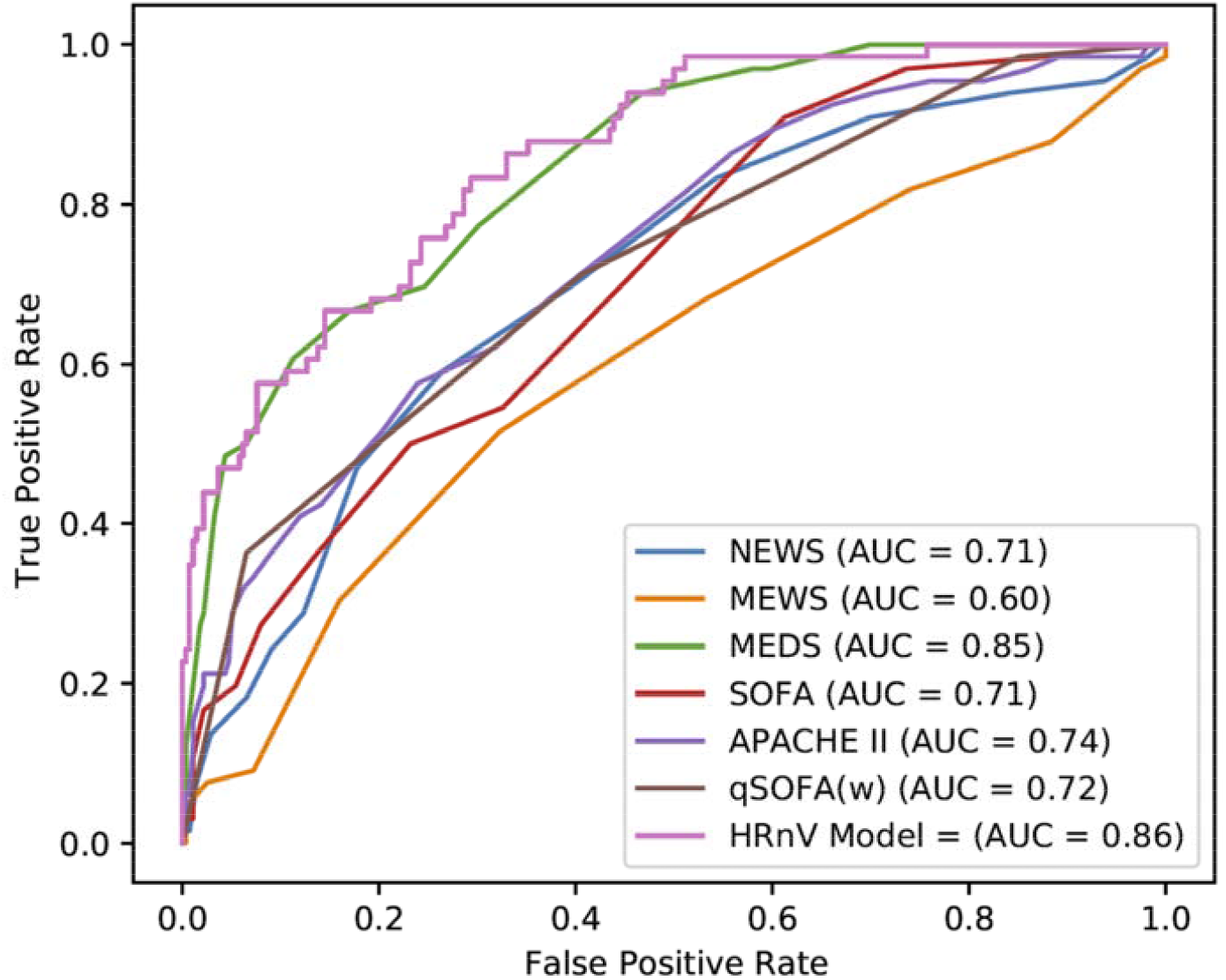
ROC curves for HRV and HRnV-based prediction models, alongside other disease severity scoring systems. Confidence intervals shown are for 95%. APACHE II: Acute Physiology and Chronic Health Evaluation II; AUC: area under the receiver operating characteristic curve; HRnV: heart rate n-variability; HRV: heart rate variability; MEDS: Mortality in Emergency Department Sepsis; MEWS: Modified Early Warning Score; NEWS: National Early Warning Score; SOFA: Sequential Organ Failure Assessment; qSOFA(w): worst quick SOFA score.

## Discussion

In recent years, there has been a surge in research interest in HRV and its ability to prognosticate for adverse patient outcomes across various disease processes^18,19,29^. To improve the predictive ability of HRV, several studies have sought to utilise advanced nonlinear techniques to derive novel HRV parameters^30,31^. Indeed, we previously employed the above novel HRnV measures to assess for risk of 30-day major adverse cardiac events (MACE) in patients presenting to the ED with chest pain to good effect^21^.

In this study, we evaluated the predictive value of novel HRnV measures (HR_*n*_V and HR_*n*_V_*m*_) in assessing the risk of 30-day IHM in patients presenting to the ED with sepsis. In addition to the 22 traditional HRV parameters, we derived an additional 120 HRnV parameters, 71 of which were found to be statistically significant in their association with the primary outcome. The newly generated HRnV parameters greatly amplify the number of candidate predictors, and have demonstrated improved predictive ability for sepsis mortality. The newly added parameters, NN50*n* and pNN50*n*, are significantly associated with mortality in the univariate analysis, and characterise the number of times that the absolute difference between two successive RR_*n*_I sequences exceeds 50×*n* ms, by assuming the absolute difference may be magnified when the corresponding RR_*n*_I is *n* times longer than RRI^21^. The composite HRV-HRnV model as derived from multivariable logistic regression achieved the highest AUC on ROC analysis, and outperformed other established disease scoring systems such as NEWS, MEWS, MEDS, SOFA, and APACHE II for the prediction of 30-day IHM in patients presenting to the ED with sepsis.

In addition to demonstrating superior predictive ability for sepsis mortality, the HRnV model is made even more relevant in its ability for rapid and objective prognostication where only vital signs and parameters calculated from five-minute ECG tracings are needed. Many established disease severity scores require invasive tests which require long turnaround times and resources to obtain or include subjective parameters that involve interrater variability while scoring. Among disease severity scores with higher AUCs, the MEDS score, developed specifically for the purpose of risk stratification of septic patients in the ED, suffers from some of these limitations and its adoption has thus not been widespread. APACHE II and SOFA scores initially designed for use in the intensive care unit (ICU) setting similarly requires invasive investigations in the calculation of its score. In these aspects, the HRnV model which is derived from vital signs taken on ED presentation, and HRV and HRnV measures calculated from a five minute ECG tracing, can overcome these limitations, and provide a rapid, objective, and accurate risk assessment of the septic patient. A triage tool with these characteristics would be invaluable to the physician and can aid in risk stratification, clinical management, patient disposition, and accurate patient classification for administrative or research purposes.

### Limitations

Despite the presentation of many HRnV parameters in this study, there exist technical limitations which constrain their utility. First, a majority of the HRnV variables were removed from predictive modelling with traditional logistic regression, which hindered the release of the power of the novel HRnV representation. Moving forward, we endeavour to explore the use of machine learning algorithms^37-39^ for full utilization of the HRnV parameters. Second, the difficulty in interpretating the HRnV parameters may pose a challenge in their clinical implementation and adoption. Third, although the novel HRnV variables and corresponding model have shown strong predictive value and potential clinical utility, this study was a single-center, retrospective observational study, thus multi-center trials would be required to corroborate our results. Prospective cohort studies would also be valuable in validating the effectiveness of the HRnV parameters, and more conclusively establishing a cause and effect relationship.

## Conclusions

The use of novel HRV measures (HR_*n*_V and HR_*n*_V_*m*_) can provide additional power to predictive models in the risk stratification of patients who present to the ED with sepsis. When included in a model with other clinical variables, the HRnV model outperforms traditional risk stratification scoring systems. The use of HRnV measures may allow for a rapid, objective, and accurate means of patient risk stratification for sepsis severity and mortality.

## Data Availability

The datasets used and/or analyzed during the current study are available from the corresponding author on reasonable request.

